# Polygenic prediction of major depressive disorder and related traits in African ancestries UK Biobank participants

**DOI:** 10.1101/2023.12.24.23300412

**Authors:** SC Kanjira, MJ Adams, Yunxuan Jiang, Chao Tian, 23andMe Research Team, CM Lewis, K Kuchenbaecker, AM McIntosh

## Abstract

**Introduction:** Genome-Wide Association Studies (GWAS) over-represent European ancestries compared to the global population, neglecting all other ancestry groups and low-income nations. Consequently, polygenic risk scores (PRS) more accurately predict complex traits in Europeans than African Ancestries groups. Very few studies have looked at the transferability of European-derived PRS for behavioural and mental health phenotypes to non-Europeans. We assessed the comparative accuracy of PRS for Major Depressive Disorder (MDD) trained on European and African Ancestries GWAS studies to predict MDD and related traits in African Ancestries participants from the UK Biobank.

**Methods:** UK Biobank participants were selected based on Principal component analysis (PCA) clustering with an African genetic similarity reference population and MDD was assessed with the Composite International Diagnostic Interview (CIDI). Polygenic Risk Scores (PRS) were computed using PRSice2 using either European or African Ancestries GWAS summary statistics.

**Results:** PRS trained on European ancestry samples (246,363 cases) predicted case control status in Africans of the UK Biobank with similar accuracies (190 cases, R^2^=2%) to PRS trained on far much smaller samples of African Ancestries participants from 23andMe, Inc. (5045 cases, R^2^=1.8%). This suggests that prediction of MDD status from Africans to Africans had greater efficiency per unit increase in the discovery sample size than prediction of MDD from Europeans to Africans. Prediction of MDD status in African UK Biobank participants using GWAS findings of causal risk factors from European ancestries was non-significant.

**Conclusion:** GWAS studies of MDD in European ancestries are an inefficient means of improving polygenic prediction accuracy in African samples.

## INTRODUCTION

Depressive disorders are ranked as the third leading cause of disability, as measured by years lived with disability, with Major Depressive Disorder (MDD) being the most significant contributor to this burden. The World Health Organization estimates that more than 322 million individuals globally suffer from MDD, with at least 9% of these cases occurring in Africa (1,2). While lower rates of MDD have been reported in Africa compared to Europe and North America, recent studies suggest that MDD is under-reported in Africa and that most affected individuals go undiagnosed (3,4).

MDD has a heritability of 30-40% (5) and better characterisation of its genetic architecture may provide both an improved mechanistic understanding and more accurate genetic prediction. So far, genome wide association studies (GWAS) have successfully identified over 243 variants to be associated with depression, focussing on participants of European ancestry (6). Sirugo et al., (2019) showed that GWAS studies over-represent European compared to other ancestry groups, with an approximately 5-fold over-representation compared to their global population (7–9). The overrepresentation of Europeans in genetics research means that the potential benefits of these studies will disproportionately apply to people of European ancestry and deprive other ancestries and low-income countries of new treatments and diagnostics (10).

Due to overrepresentation of Europeans in GWAS, polygenic risk scores (PRS) developed from these studies more accurately predict many complex traits in European than in African Ancestries samples (11,12). The difference in prediction may be due to differences in the phenotypes themselves, their genetic architectures or because of gene-by-environment interactions (13–15). Very few studies have looked at the transferability of European-derived PRS for behavioural and mental health phenotypes to non-Europeans generally and Africans specifically. Consequently, the predictive accuracy of European derived MDD-PRS to African samples remains uncertain. We looked at the transferability of MDD-PRS trained on European GWAS studies to African Ancestries participants from the UK Biobank within and across traits. Furthermore, we sought to compare the transferability of MDD-PRS trained in participants of African ancestries from 23andMe Inc., (mainly from North America), with the transferability of MDD-PRS trained on Europeans to the African-ancestry participants from the UK Biobank.

## METHODOLOGY

### SAMPLES

The study focused on participants in the UK Biobank who are genetically similar to 1000 Genomes African reference samples. The UK Biobank is a prospective cohort study of over 500 000 individuals of diverse ethnic backgrounds from across the United Kingdom (16). These participants volunteered to be part of the UK Biobank between 2006 and 2010 and submitted saliva, blood, urine, physical measurements. Participants also completed questionnaires and a verbal interview and later this data was linked to health records such as primary care, hospital in-patient records, cancer register data, and death records. An online Mental Health Questionnaire that included a depression assessment was sent to participants by email and entitled ‘The thoughts and feelings questionnaire’ (17). The questionnaire was offered to the 317 785 participants, out of the total 502 616 UK Biobank participants, who had agreed to email contact, and 157 396 completed the online questionnaires by June 2018.

A depression phenotype was generated based on the CIDI-SF (Composite International Diagnostic Interview Short Form) (18). Cases were defined as those participants who had at least one core symptom of depression (persistent sadness or loss of interest) for most of the day or all of the day. Symptoms had to be present for a period of over two weeks plus another four non-core depressive symptoms that represent a change from usual occurring over the same timescale, with some or a lot of impairment. Cases that self-reported another mood disorder were excluded. Controls were defined as participants who did not meet symptom criteria for MDD (17,19).

### African Ancestries participants in UK Biobank

Principal component analysis (PCA) was used to select people of African ancestries from the UK Biobank. First, we selected individuals who self-reported as being Black or Black British (Caribbean, African, Any other Black background), White and Black Caribbean or Black African, and participants whose self-identity was not specifically categorised (responses "Other ethnic group", "Any other mixed background", "Do not know", or "Prefer not to answer’). Using the genotypes provided by UK Biobank, we derived ancestry informative genetic principal components using the weights from the 1000 Genomes reference dataset to cluster the participants into their genetic similarity groupings. UK Biobank participants who were similar to the 1000 Genomes African (AFR) cluster were then selected for further analysis.

#### Polygenic risk scores

Polygenic risk scores were computed using PRSice-2 software (20) using the clumping and thresholding method (C+T) so that only SNPs that are weakly correlated with one another are retained (20). After clumping, SNPs with a p value larger than a specified level of significance were removed, PRS were then calculated by the sum of SNP allele effect sizes multiplied by the number of risk alleles. Both the base and target data sets were quality controlled (QC) by removing ambiguous and duplicate SNPs, SNPs with a minor allele frequency (MAF) of less than 1% and a genotype missingness greater than 2% were also removed. For every trait, we report empirical P values after specifying 10 000 permutations in PRSice-2.

#### Summary statistics

To compute polygenic risk scores in African-clustered participants of the UK Biobank, we used GWAS summary statistics of MDD from global European studies (246 363 cases and 561 190 controls), an African American study from 23andMe (5045 cases, 102 098 controls), a secondary dataset comprising summary statistics from a meta-analysis of 12 African cohorts (36 313 cases, 160 775 control) and data from several traits that have been known to be associated with depression from European-clustered studies, in addition to height, which we used as a negative control. For each set of summary statistics, SNP based heritability was calculated using linkage disequilibrium score regression as implemented in the LDSC software package (21,22).

Table 1 above details the summary statistics used in PRS analysis, detailing source, ancestry group, number of cases and controls, and SNP-based Heritability for each study. Heritability was calculated using the LDSC (LD Score) package.

**Table 1:** Summary Statistics Used in PRS Analysis.

| **Phenotype** | **Ancestry group** | **N Cases** | **N controls** | **$h^2_{\text{SNP}}$ (%)** | **Reference** |
| --- | --- | --- | --- | --- | --- |
| Major depression | European | 246 363 | 561 190 | 6.68 | (Howard et al., 2019), (23) |
| Major depression | African American | 5045 | 102 098 | -0.72 | 23andMe, unpublished |
| Major depression | African | 36 313 | 160 775 | 2.77 | Meng et al., 2022 (unpublished) |
| BMI | European | 339 224 | — | 12.21 | (Locke et al., 2015), (24) |
| Height | European | 4 080 687 | — | 38.47 | (Yengo et al., 2022), (25) |
| Schizophrenia | European | 67 390 | 94 015 | 16.22 | (Trubetskoy et al., 2022), (26) |
| Educational attainment | European | 765 283 | — | 12.11 | (Okbay et al., 2022), (27) |
| Bipolar Disorder | European | 41 917 | 371 549 | 9.86 | (Mullins et al., 2021), (28) |
| Neuroticism | European | 63 661 | — | 2.71 | (de Moor et al., 2015), (29) |

## RESULTS

### African-ancestries clustered participants in the UK Biobank

We utilized principal component analysis (PCA) to project the genetic data of UK Biobank participants onto the PCA space defined by the reference 1000 Genomes dataset. This approach enabled the selection of individuals from the UK Biobank whose genetic profiles closely resemble those of the African ancestry samples within the 1000 Genomes dataset. Figure 1 below is a PCA plot showing the clustering of individuals of possible African ancestry (people who self-reported as being Black or Black British, Other ethnic grouping, do not know, mixed, and prefer not to answer) in the UK Biobank with the reference 1000 Genomes dataset.

**Figure 1:**
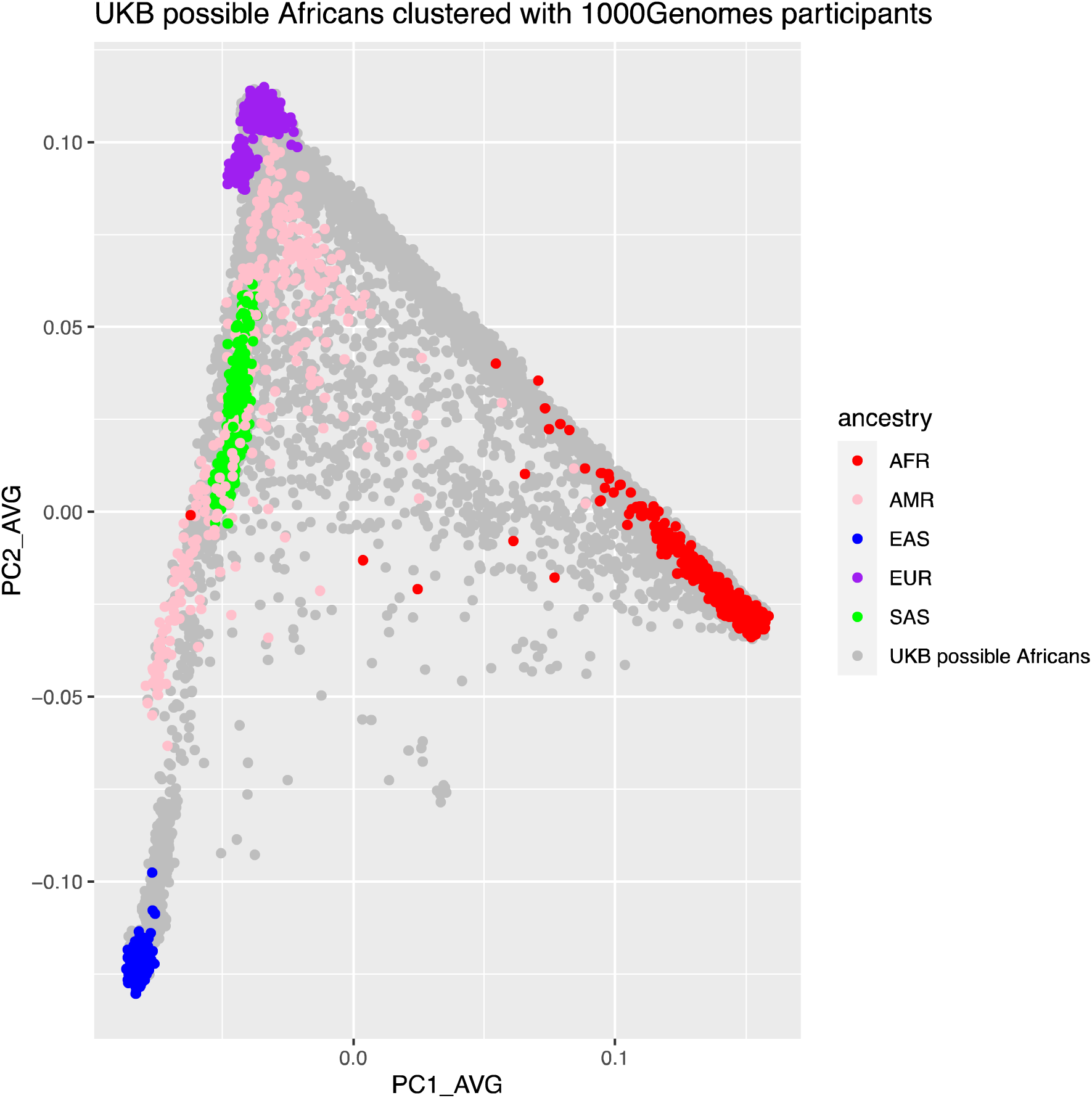
PCA plot of UK Biobank participants of self-reported African, mixed, or unknown ancestry clustered with 1000 Genomes reference data set. The different colours represent the different ancestry grouping in the reference 1000 Genomes dataset as shown in the key (AFR=Africans, AMR=Americans, EAS=Asians, EUR=Europeans and SAS=South Asians), while the grey colour show individuals of possible African Ancestries based on self-report from the UK Biobank.

UK Biobank participants of possible African Ancestries that clustered within 6 standard deviations of the 1000 Genomes AFR cluster were selected to represent Africans in the UK Biobank. In total 8543 participants in the UK Biobank clustered with the 1000 Genomes AFR reference group as shown in figure 2. Of these, 1090 participants completed the Mental Health Questionnaire, with 190 participants meeting the CIDI criteria for MDD cases and 671 participants met the criteria for controls.

**Figure 2:**
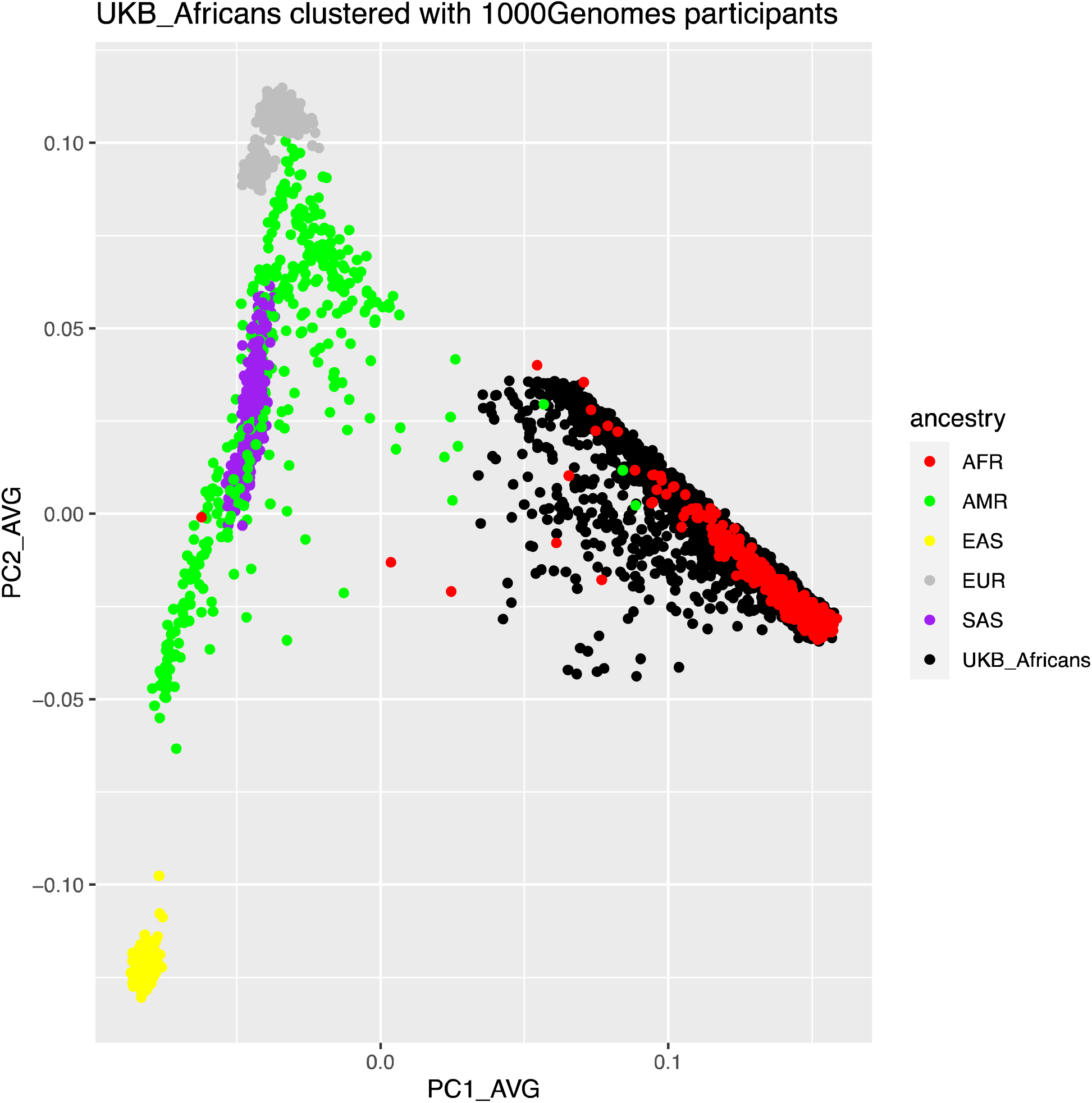
PCA plot showing UK Biobank participants (black) who clustered with the 1000 Genomes AFR (red) reference group.

### Polygenic Risk Scores

#### Within ancestry within trait polygenic prediction of MDD from African datasets to African participants in the UK Biobank

MDD GWAS results of African American participants from 23andMe (5045 cases and 102 098 controls) were used to predict MDD status in African participants of the UK Biobank. Figure 3 below indicates that the African American summary statistics significantly predicted MDD status in UK Biobank African Ancestries participants across all p-value thresholds, and the most predictive p-value threshold was 0.2 explaining 1.8% of variation in depression liability (empirical p-value = 0.008). This PRS prediction of MDD from African sample to African sample is comparable in accuracy with prediction of PRS trained on European ancestry samples of over 800K individuals (246 363 cases and 561 190 controls).

**Figure 3:**
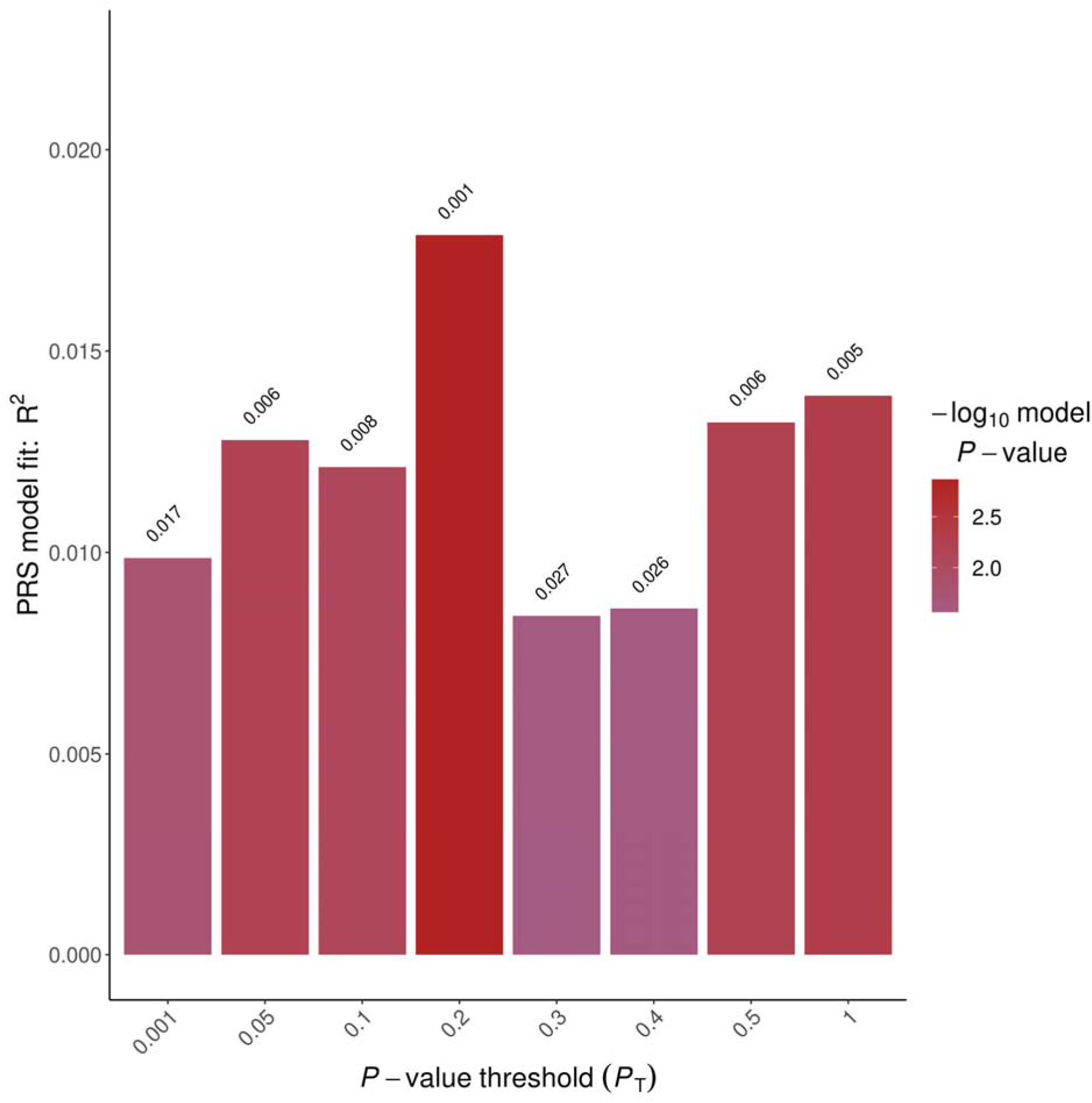
Prediction of MDD in African participants of the UK Biobank using African American data (5045 cases and 102 098 controls) from 23andMe. The y-axis displays the R2 values corresponding to the PRS predictions, while the x-axis illustrates the spectrum of p-value thresholds utilized in the PRS analysis. The p-values for the predictions are depicted atop the bars.

We also used a secondary set of summary statistics from a meta-analysed data of 12 African cohorts with 36,313 MDD cases and 160 775 controls to predict MDD status in African participants of the UK Biobank. 99.6% of the MDD cases in this meta-analysed dataset are multiple African American studies and 0.4% participants are from South Africa. In contrast, PRS trained on the meta-analysed predominantly African American dataset *did not* significantly predict MDD status in Africans of the UK Biobank as depicted in Figure 4.

**Figure 4:**
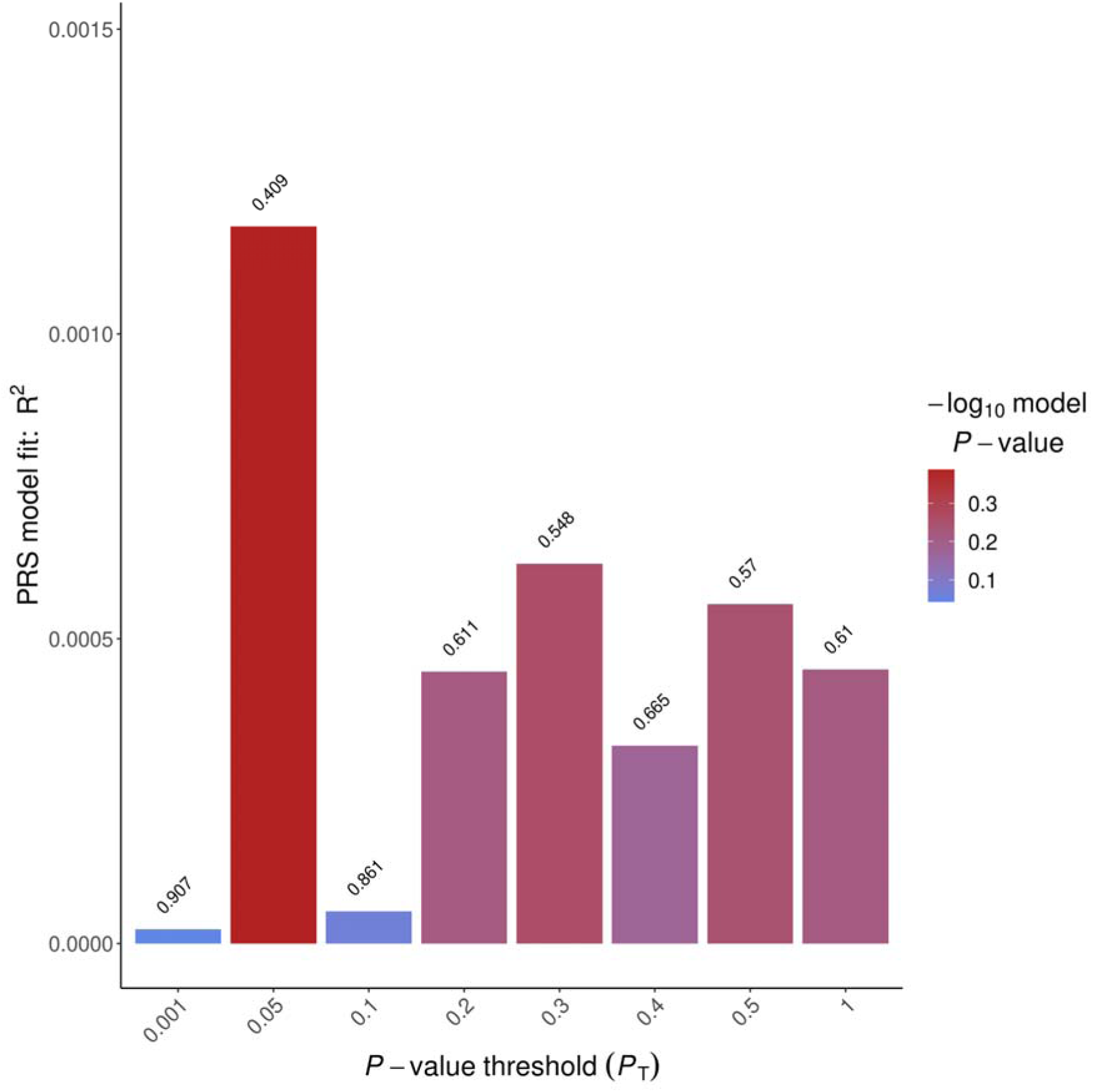
Prediction of MDD status in African participants of UK Biobank using meta-analysed African MDD summary statistics from multiple African American (99%) and one Africa-based study.

#### Cross ancestry within trait prediction of MDD and other traits

European-based GWAS results for MDD, BMI, Neuroticism, education attainment and height were used to predict within the same trait in UK Biobank African Ancestries participants. European-based GWAS results significantly predicted MDD, BMI, education attainment, and height within trait in African participants of the UK Biobank as illustrated in Figure 5.

**Figure 5:**
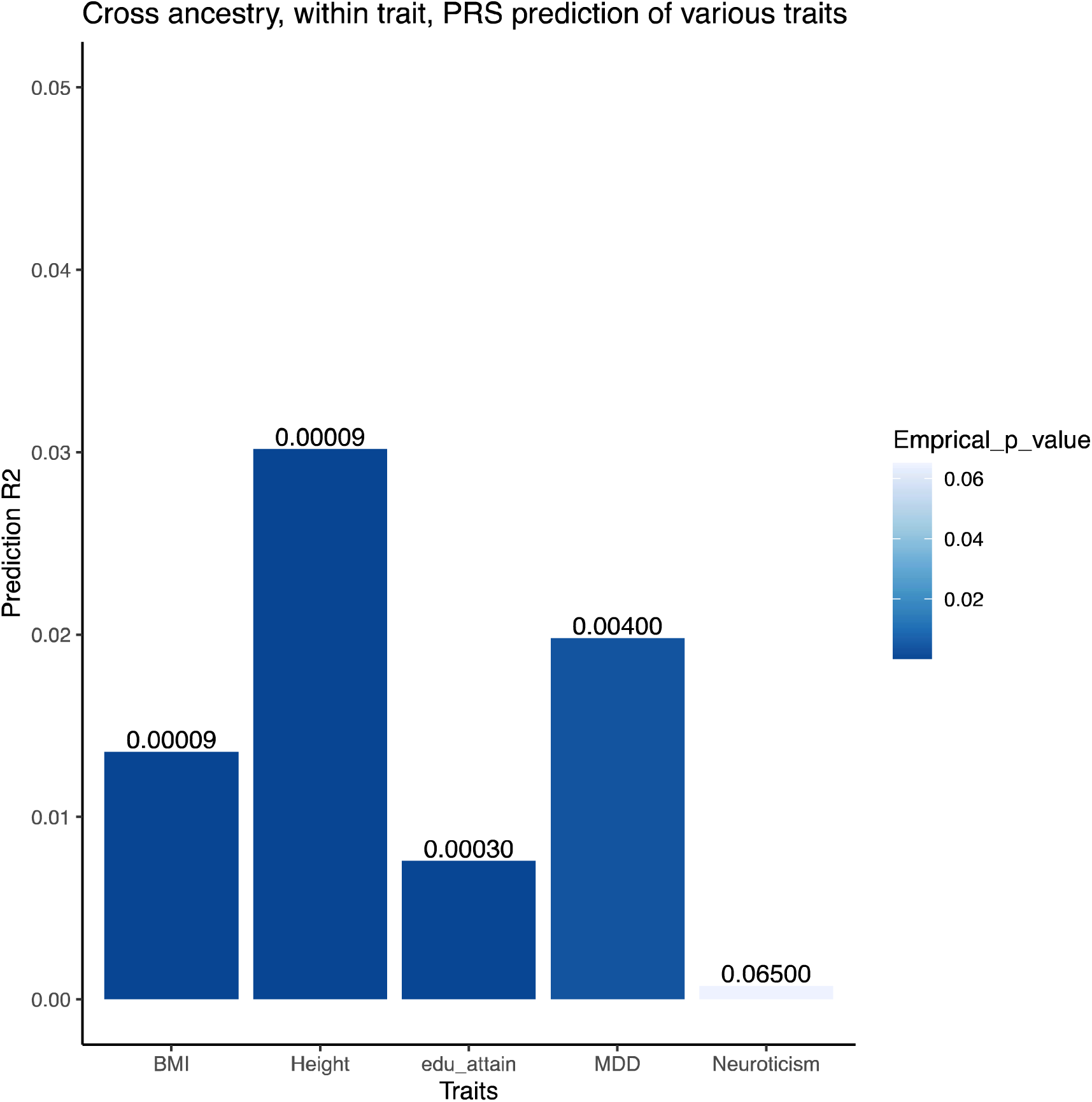
Bar plots showing cross-ancestry within trait polygenic prediction of BMI, height, education attainment (edu_attain), MDD and Neuroticism. The y-axis is showing the R^2^ values for the PRS predictions while the x-axis is showing the most predictive bar for each trait with empirical P values for the prediction shown on top of the bars.

Specifically, the European based MDD-PRS explained a 2% variation in MDD risk among individuals of African Ancestries in the UK Biobank, with an empirical P value of 0.0039. The PRS associated with education attainment explained 0.7% of the variation in education attainment within the same cohort, yielding a P value of 3×10^-4^ (figure 5). However, it is noteworthy that European-based Neuroticism PRS did not significantly predict Neuroticism in African Ancestries participants. Height PRS, a highly heritable trait used for comparison purposes, explained 3% of the variation in height among UK Biobank Africans

#### Cross ancestry cross trait polygenic prediction of MDD

Cross ancestry cross trait polygenic prediction of MDD in Africans of the UK Biobank using PRS estimated from European based GWAS summary statistics for traits known to be associated with MDD (namely: Bipolar Disorder, BMI, Schizophrenia, Neuroticism, and education attainment) did not yield any significant results as shown in figure 6. Height summary statistics were used for comparison purposes.

**Figure 6:**
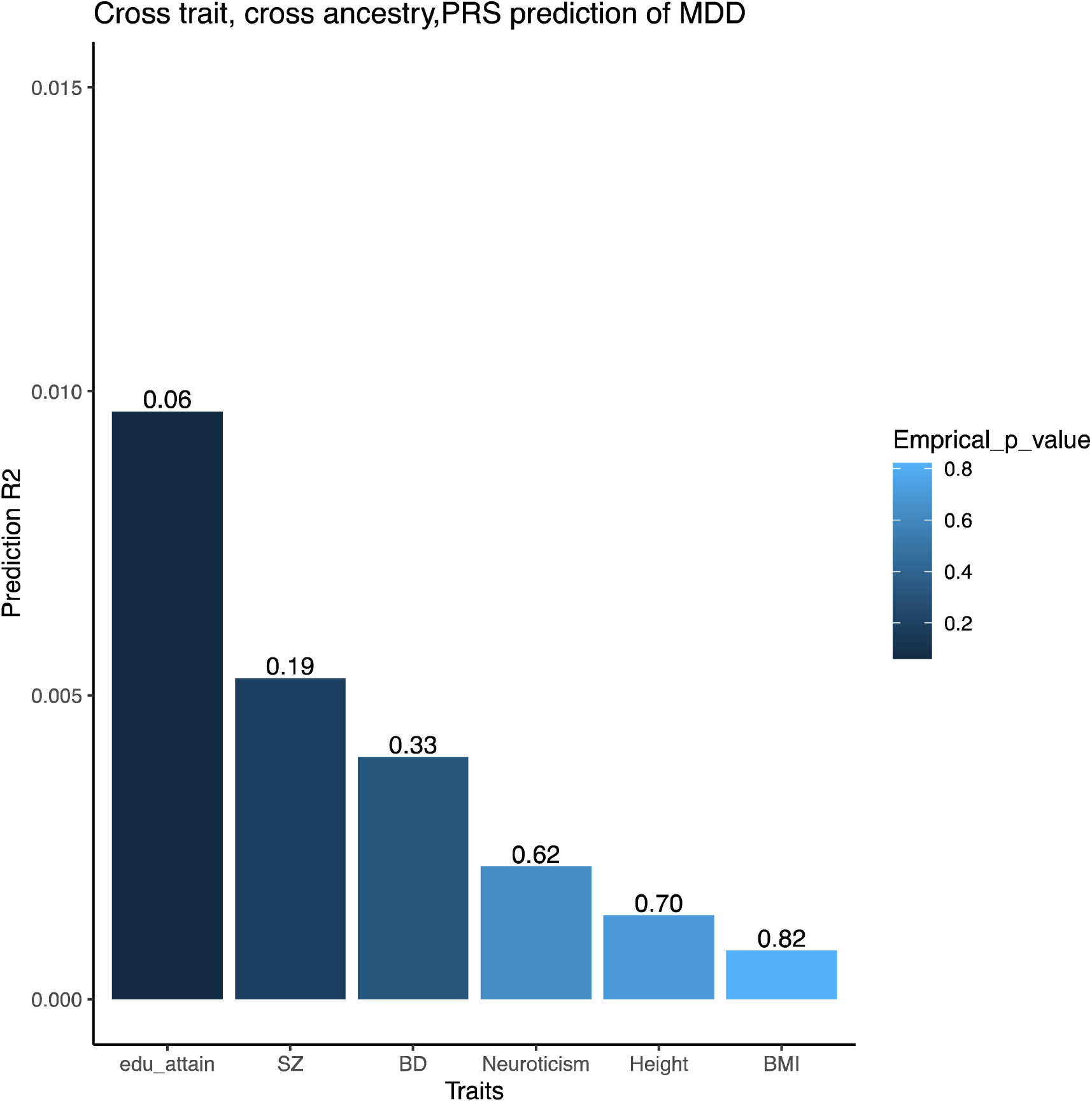
Bar plot showing cross ancestry cross trait polygenic prediction of MDD using Education attainment (edu_attain), Schizophrenia (SZ), bipolar disorder (BD), Neuroticism, Height, and BMI. The y-axis displays the R2 values for the PRS predictions, while the x-axis represents the most predictive bar associated with each trait, and the empirical P values for the predictions are presented atop these bars.

## DISCUSSION

Polygenic risk scores trained on European GWAS studies of 246 363 depression cases predicted MDD case-control status in African Ancestries participants from UK Biobank at lower prediction accuracies (R^2^=2%) than for European participants (R^2^=3.2%). Polygenic risk scores trained on a much smaller single GWAS of 5045 depression cases predicted depressed case-control status in African Ancestries participants from UK Biobank at a similar accuracy (R^2^=1.8%), suggesting a much greater prediction accuracy relative to sample from African than from European GWAS training datasets. Prediction of MDD status in African UK Biobank participants using European ancestry GWAS studies of other traits (known to predict MDD status in European populations) was non-significant. GWAS studies of depression from European samples are an inefficient means of improving polygenic prediction accuracy in African samples and genetic scores derived from European ancestry studies of known risk factors for depression may also be less useful for mechanistic studies.

In contrast to the findings made using 23andMe summary statistics, polygenic scores derived from a GWAS meta-analysis of several African ancestry studies within and outside of Africa, showed limited predictive ability for Major Depressive Disorder (MDD) in African Ancestries UK Biobank participants. The dataset combined 36 313 MDD cases and 160 775 controls, predominantly comprising African Americans, with a small representation (139 cases and 346 controls) from continental African populations (Drakenstein Child Health Study). Despite expectations of superior performance compared to the 23andMe dataset, various factors may have contributed to this underperformance. Firstly, while 23andMe used a single definition of MDD and a single genotyping quality-control pipeline, MDD phenotype definitions and methods varied across the included cohorts in the GWAS meta-analysis. Some studies employed stringent criteria while others used broader definitions. The inclusion of individuals with varying definitions of African Ancestries may also have increased genetic heterogeneity. The meta-analysed data primarily featured African Americans, who exhibit varying degrees of genetic admixture with other ancestral backgrounds, possibly influencing the accuracy of PRS predictions in UK Biobank. A recent study by Ding et al. in 2023 revealed that for highly polygenic traits, PRS predictive accuracy tends to diminish with increasing genetic distance between populations (30).

Several European ancestry studies have shown that various traits have a shared genetic liability with MDD, some of which may be causally associated but little is known about the shared genetic liability of MDD with other traits across ancestries (31). We looked at cross ancestry cross traits prediction of MDD using height, BMI, bipolar disorder, schizophrenia, neuroticism, and education attainment in people of African Ancestries using European based GWAS results. Height was used as a highly heritable control trait with no known causal relationship with MDD. While our study did not yield successful predictions of MDD status in Africans of the UK Biobank using European GWAS results of various traits, it is worth noting that previous investigations conducted within European populations have demonstrated a shared genetic liability between MDD and traits such as Bipolar Disorder, BMI, and neuroticism (32–34). To advance our understanding of shared genetic liability in African populations, future research endeavours could explore this aspect by training PRS using GWAS data derived specifically from African cohorts. This approach has the potential to uncover novel insights into the shared genetic components between MDD and other traits within the context of African ancestral backgrounds.

The predictive accuracy of MDD-PRS trained on African American data from 23andMe was comparable to that of PRS trained on European data among individuals of African Ancestries, despite the African American PRS being based on a considerably smaller sample size compared to the European PRS. This observation aligns with findings from other studies focused on various traits, suggesting that PRS derived from African Ancestries data tends to exhibit superior performance when applied to African populations than PRS derived from European data.

## Data Availability

All data produced in the present study are available upon reasonable request to the authors

## Acknowledgements

This work was supported by UK Medical Research Council Grant (reference MR/S035818/1), Wellcome Trust grants (references 223165/Z/21/Z [DepGenAfrica], 220857/Z/20/Z [WTIA]), the European Union H2020 Scheme (under action Grant agreement 847776) and by the US National Institutes of Health (1R01MH124873-01). We would like to thank the African ancestries research participants of UK Biobank and all of the studies used for the work presented here. We would also like to thank the research participants and employees of 23andMe, Inc. For making this work possible.

The following members of the 23andMe Research Team contributed to this study: Stella Aslibekyan, Adam Auton, Elizabeth Babalola, Robert K. Bell, Jessica Bielenberg, Jonathan Bowes, Katarzyna Bryc, Ninad S. Chaudhary, Daniella Coker, Sayantan Das, Emily DelloRusso, Sarah L. Elson, Nicholas Eriksson, Teresa Filshtein, Pierre Fontanillas, Will Freyman, Zach Fuller, Chris German, Julie M. Granka, Karl Heilbron, Alejandro Hernandez, Barry Hicks, David A. Hinds, Ethan M. Jewett, Yunxuan Jiang, Katelyn Kukar, Alan Kwong, Yanyu Liang, Keng-Han Lin, Bianca A. Llamas, Matthew H. McIntyre, Steven J. Micheletti, Meghan E. Moreno, Priyanka Nandakumar, Dominique T. Nguyen, Jared O’Connell, Aaron A. Petrakovitz, G. David Poznik, Alexandra Reynoso, Shubham Saini, Morgan Schumacher, Leah Selcer, Anjali J. Shastri, Janie F. Shelton, Jingchunzi Shi, Suyash Shringarpure, Qiaojuan Jane Su, Susana A. Tat, Vinh Tran, Joyce Y. Tung, Xin Wang, Wei Wang, Catherine H. Weldon, Peter Wilton, Corinna D. Wong.

Cathryn Lewis is part-funded by the NIHR Maudsley Biomedical Research Centre at South London and Maudsley NHS Foundation Trust and King’s College London. The views expressed are those of the author(s) and not necessarily those of the NIHR or the Department of Health and Social Care.

## CONFLICT OF INTEREST

Yunxuan Jiang, Chao Tian are employed by and hold stock or stock options in 23andMe, Inc. All other authors declare no conflict of interests.

Cathryn Lewis sits on the scientific advisory board for Myriad Neuroscience, has received speaker fees from SYNLAB, and consultancy fees from UCB.

## Notes

### Competing Interest Statement

The authors Yunxuan Jiang, Chao Tian are employed by and hold stock or stock options in 23andMe, Inc., All other authors declare no conflict of interests.
Cathryn M Lewis sits on the scientific advisory board for Myriad Neuroscience, has received speaker fees from SYNLAB, and consultancy fees from UCB.

### Author Declarations

The North West Multi-Centre Research ethics Committee of The National Health Service gave ethical approval for this work.

